# Relevance-Based Prediction: A Transparent, Patient-Specific Approach to Predicting Medical Outcomes

**DOI:** 10.64898/2026.09.28.26364225

**Authors:** Christopher L. Robinson, David Turkington, Linda Lee, Mark Kritzman, R. Jason Yong

## Abstract

Accurate prediction of individual medical outcomes is essential for optimizing treatment allocation amid rising costs, coverage denials, and limited clinical resources. Traditional predictive models, including linear regression and neural networks, rely on average effects and cannot tailor predictions to the specific circumstances of individual cases. We present relevance-based prediction (RBP), a model-free and task-specific method that predicts outcomes as weighted averages of observed cases, with weights determined by a rigorously defined and theoretically justified measure of relevance. Unlike model-based methods that apply an estimated relationship to patient-specific characteristics to capture conditional effects, RBP revisits the original data for each prediction and measures the alignment of the relevance weights and outcomes for many combinations of cases and variables. In this way RBP explicitly considers which cases and variables contribute to each specific prediction as well as the strength of the patterns in the data that support that prediction. We illustrate RBP by applying it to predict opioid treatment outcomes and demonstrate how it provides case-specific insights that build on the insights given by conventional models, including how each historical case contributes to a specific prediction, how each variable affects each predicted outcome and the reliability of that prediction, and how reliably each outcome can be predicted before it is known. These additional insights may help to prevent misleading average-based decisions and reduce harmful or suboptimal treatment.

## Introduction

Health care expenditures are escalating worldwide. Medical costs are projected to increase globally by ~10.3%^1,2^ In the USA, healthcare spending totaled over US$5.3 trillion in 2024, averaging to US$15,474 per person, accounting for ~18.0% of GDP and rising over 7.2% from the previous year, surpassing GDP growth.^3,4^ Resources are continuing to be overstrained: hospital care, physician and clinical services, and prescription drugs together consume a significant share of that expenditure, and demand for treatments is rising.^5–7^ Alongside the financial pressures, prediction of medical outcomes, such as response to treatment, risk of complications, or long-term morbidity, remains limited. Predictive performance can vary substantially across patient populations and clinical settings, and established tools may perform less well in patients with multimorbidity.^8,9^ Models that inadequately represent interactions between treatment and patient characteristics may also give misleading estimates of treatment benefit for individuals.^10^ The inability to forecast which patients are most likely to benefit (or to be harmed) leads to inefficient use of interventions, over-treatment, and waste, as well as undertreatment in high-risk groups. There is a pressing need for more accurate, validated prediction models in treatment response and outcome trajectories, which would allow health systems to better direct scarce resources (*e.g*., costly therapies, specialist visits, hospital beds, etc.) toward patients most likely to derive benefit, improving both effectiveness and equity while restraining costs.

Despite the promise of predictive modeling in medicine, especially with machine learning, its real-world performance has often fallen short, illustrating the risks and opportunity costs of imprecise prediction. In migraine management, a recent systematic review and meta-analysis found that prediction models had a pooled area under the curve of 0.86 (95% CI, 0.67–0.95), yet most studies carried high risk of bias and substantial heterogeneity, limiting confidence in clinical deployment.^11^ More broadly, clinical risk models often suffer from instability and miscalibration when applied to new populations. One analysis demonstrated that many models’ predictions vary greatly when retrained on bootstrapped subsets, thus undermining robust performance in external settings.^12^ Furthermore, embedding predictive models into scarce-resource allocation frameworks (*e.g*., for organ transplantation) has in some cases exacerbated inequities, because biases in the training data propagate into unfair prioritization across demographic groups.^13,14^ On the other hand, novel predictive strategies applied to resource planning (*e.g*., dynamic model predictive control tools to allocate interventions iteratively) demonstrate potential in simulation and early implementation, enabling responsive and optimized deployment of limited capacity.^15,16^ Better predictive models could enable health systems to direct high-cost therapies, advanced diagnostics, and vigilant monitoring toward patients with the greatest likelihood of benefit, thereby maximizing outcomes under constrained budgets.

A critical obstacle to effective prediction of medical outcomes is the limited transparency of model-based approaches to prediction. Models are mathematical prediction rules that contain parameters whose values must be estimated from a training sample. After the parameter values are computed the model makes predictions based on these values and no longer requires the data. The data is typically discarded and is not available to a practitioner who relies on the model’s predictions for guidance. Moreover, a model’s parameter values are chosen to minimize the average prediction error across the training sample. It is difficult, if not impossible, for a practitioner who can only observe parameter values and associated statistical tests to ascertain the prevalence and characteristics of prior cases that inform a specific prediction. For example, a prediction might be influenced heavily by a prior case for which there is a confounding condition that is not captured by the model. If one is made aware of this case-specific dependence, he or she may be inclined to investigate this case, decide whether to remove it or retain it in the case sample, and judge confidence in the prediction accordingly.

Additionally, most model interpretability methods focus on the average influence of predictive variables on the reliability and magnitude of a prediction. A regression model’s p-value estimates how a predictive variable affects the average reliability across all predictions, and its β coefficients reflect average impact on the prediction value across a wide range of predictions. Moreover, these metrics measure marginal effects, which could be distorted by collinearity. The SHAP method, which is commonly used to measure variable importance in machine learning models, does measure total effects rather than marginal effects, but its assessment of a variable’s contribution to the reliability of a prediction is based on the model’s parameters. Therefore, it cannot evaluate how the variables impact the reliability of specific predictions in the context of how many observations informed that prediction from the training sample.^17^ Model-based prediction may fail to detect that a variable, though having a positive influence on average, could be harmful to specific patients. Finally, model-based summary statistics reveal the average reliability of predictions. For example, a linear regression’s R-squared measures the average predictive efficacy of the model. It is based on some good predictions, some mediocre predictions, and some bad predictions. It does not necessarily reveal much about the trustworthiness of a specific prediction. Likewise, the mean squared errors (MSE) of a machine learning model’s predictions for a full or partial sample of training data reflects an average over many circumstances rather than an assessment tailored to one specific prediction. In these cases, the reliability of a specific prediction is most directly revealed after one observes the outcome.

We describe for the first time within a medical context an alternative prediction method called relevance-based prediction (RBP), which has previously been introduced to finance and sports analytics.^18–20^ RBP gives full visibility into the formation of each prediction, and it gives advance guidance of each prediction’s reliability. We show in the appendix that fit is an exact decomposition of R-Squared in linear regression. The enhanced visibility that RBP gives into the formation of individual patient predictions may guide a caregiver to choose a different treatment plan for a patient whose predicted outcome is known in advance to be untrustworthy compared to a patient whose outcome can be confidently predicted.

Specifically, RBP provides the following information.

1. RBP shows explicitly how each case in the sample informs each prediction, thereby enabling researchers to discount a prediction or to boost their confidence in a prediction because of its dependence on certain cases. This detailed information may lead researchers to adjust the composition of the case sample to derive a more reliable prediction.
2. RBP shows how each predictive variable impacts the total reliability and total value of each prediction, which could lead caregivers to tailor treatment in accordance with each patient’s unique circumstances.
3. RBP gives advance guidance about each prediction’s reliability in two separate but complementary ways. First, it quantifies the extent to which there are useful patterns in a dataset for forming a prediction by measuring the alignment of the observation weights and the outcomes for each prediction. This alignment captures the strength of the signal in the data. Second, it shows the distribution of the information that goes into each prediction, which captures the extent to which the cases used to form the prediction agree or disagree. This distribution of information captures the noise around the signal.^21^

We first describe RBP conceptually and mathematically. We then illustrate it by predicting impact scores of patients who have been treated with opioids using a comprehensive dataset of more than 5,800 cases; however, we wish to stress that RBP is completely general and can be applied to any data-driven prediction task. We conclude with a discussion. The descriptions of RBP and other prediction methods described herein follow closely from previously published articles and books with modifications to suit the context of the current discussion.^18–25^

### Relevance-Based Prediction: How It Works

RBP forms a prediction as a weighted average of observed outcomes in which the weights are based on a rigorously defined and theoretically justified statistic called relevance. It also depends on a statistic called fit which measures the strength of the patterns in the data that is used to form each prediction. Fit gives advance guidance about the efficacy of the cases and variables used to form each prediction. Its third feature is called grid prediction, which gives RBP its power. It enables consideration of many combinations of cases and predictive variables to form a reliability-weighted composite prediction for each prediction task.

#### Relevance

Relevance measures the importance of a previously observed case to the current prediction task based on a set of variables that describe both circumstances. Relevance is composed of similarity and informativeness, which are both measured using a statistic called the Mahalanobis distance (MD).^26^ The MD is an elegant measure of the distance between values of a set of variables that accounts not only for how the corresponding values compare independently but how they interact with each other. Similarity measures the multivariate nearness of cases to each other. Informativeness measures the multivariate distance of cases from average and therefore the unusualness of cases. An important tenet of information theory is that unusual cases warrant close attention because they contain more information than typical cases.^27^ The bottom line is that cases that are like the current case but different from the average of all cases are more relevant than those that are not.

If there were no relevance values, we would be limited to forming a prediction as an equally weighted average of the outcomes of prior cases which yields a static prediction that does not explain any variation in outcomes. With relevance, however, we can increase or decrease the weight of each prior outcome according to its relevance. These predictions account for the unique circumstances of each prediction task. It is important to note that RBP converges to linear regression under special circumstances. When relevance weights are applied to every case in a sample using the full set of predictive variables, the weighted average of outcomes gives exactly the same prediction as linear regression.^18^ From this equivalence, it is clear to see that the commonly used approach of linear regression exhibits both desirable and undesirable behaviors. It places large positive weights on relevant cases, implying that similar outcomes may occur, which makes sense. However, it places large negative weights on the least relevant cases, reasoning that the opposite of the most opposite experiences will occur. Predicting the opposite-of-opposites only works for relationships that are stable and symmetric, which does not always hold. RBP is based on the premise that we can form a better prediction by focusing on the most informative and reliable subsamples of relevant cases. RBP uses the MD to measure a case’s similarity to the current case and the informativeness of the other cases and the current case. The MD is given by equation 1.

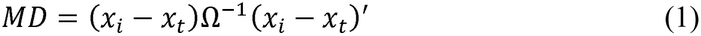

In equation 1, *x*_*i*_ is a row vector of the values of the predictive variables for a previous case, *x*_*t*_ is a row vector of the values of the predictive variables for the current case, Ω^−1^ is the inverse of the covariance matrix of the values of the predictive variables for all cases in the sample, and ′ denotes matrix transpose which converts a row vector to a column vector. The vector of the differences between *x*_*i*_ and *x*_*t*_ measures how different the values of the predictive variables for a previous case are from their values for the current case independently. By multiplying this difference vector by the inverse of the covariance matrix, we account for the covariation of the variables of the previous cases. Also, this calculation standardizes the differences by dividing them by variance. By multiplying the product by the transpose of the vector differences we consolidate the outcome into a single number, which represents the covariance-adjusted distance between the two vectors. We next show how the MD is used to determine the relevance of a case to a prediction.

The relevance of a previous case to a prediction is shown by equations 2 through 5. It is equal to the sum of its similarity to the current case and the average of its informativeness and the informativeness of the current case. We include the informativeness of the current case to center relevance on zero. Whether we include it does not affect the prediction.

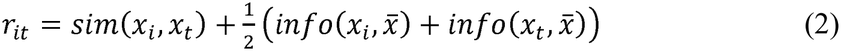

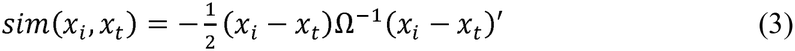

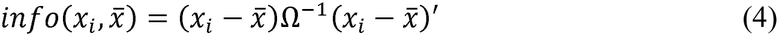

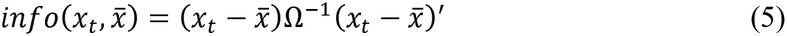

Equation 3 gives the similarity of a previous case to the current case. It is the same as equation 1 except that it is multiplied by −1/2. Because the MD measures distance, its negative value gives a measure of nearness, which is what we want to assess the similarity of one case to another. We multiply by 1/2 because the average squared distance between cases is twice as large as the average squared distance of the cases from their average. By multiplying by 1/2 we put similarity on the same scale as informativeness which is measured as a case’s Mahalanobis distance from average. Equation 4 gives the informativeness of a previous case whereas equation 5 gives the informativeness of the current case. Since equations 4 and 5 measure the Mahalanobis distance from the average values of the cases, they capture their unusualness and therefore their informativeness.

As noted earlier, RBP rests on the premise that we may be able to form more reliable predictions by using subsamples of relevant cases rather than the full sample of cases which may include non-relevant cases that obscure important patterns in the data. We use a statistic called fit to determine the most effective subsamples of relevant cases.

### Fit

Before the subsequent outcome is known, fit measures the extent to which there are useful patterns in the dataset to support a given prediction task. To the extent there is stronger alignment between the prediction weights and the outcomes, we should expect a more reliable prediction. Although other methods may provide advance guidance about the quality of individual predictions, they often rely on model-based estimates of uncertainty. RBP assesses reliability based on the empirical relationships that are relevant to each specific prediction. The classical R-squared statistic provides information about a model’s quality, but it is a grand average. By contrast, fit gives advance notice of the quality of each individual prediction, distinct from the next. It is analogous to an R-squared for an individual prediction. In fact, we have proven that fit is a mathematically exact decomposition of R-squared in the case of linear regression. We give a proof of this result in the appendix.

To see how fit is computed, consider a pair of cases that is used to form a prediction. Each case has a relevance weight and an outcome. We are interested in the alignment of the weights of the two cases with their outcomes. We first standardize them by subtracting the average value and dividing this difference by standard deviation – in essence, converting them to z-scores. We then measure their alignment by taking the product of these standardized values. If the product is positive, their relevance is aligned with their outcomes, and the larger the product, the stronger the alignment. We perform this calculation for every pair of cases in our sample. We should also note that all the formulas we have thus far considered for weights rely only on relevance, which in turn relies only on the *x*_*i*_s, the *x*_*t*_, and the 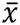. They do not use any of the information from observed outcomes. To determine fit, however, we must consider outcomes (the *y*_*i*_s) as seen in equation 6.

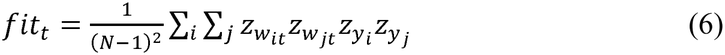

Although we compute fit from the full sample of cases, the weights that determine fit vary with the threshold we choose to define the relevant subsample. As we focus the subsample on cases that are more relevant, we should expect the fit of the subsample to increase, but we should also expect more noise as we shrink the number of cases. The fit across pairs of all cases in the full sample implicitly captures this tradeoff between subsample fit and noise by overweighting cases that are more relevant and underweighting cases that are less relevant.

Censoring cases that fall below a relevance threshold is more effective to the extent there is asymmetry between the fit of the weights formed from the retained subsample of cases and the fit of the weights formed from the complementary sample of censored cases. We measure asymmetry between the fit of the retained and censored subsamples as shown by equation 7. The (+) superscript designates weights formed from the retained cases while the (−) superscript designates weights formed from the censored cases. Asymmetry recognizes the benefit of censoring non-relevant cases that contradict the predictive relationships that exist among the relevant cases. This assessment also inherently considers the relative sizes of the two subsamples.s assessment also inherently considers the relative sizes of the two subsamples.

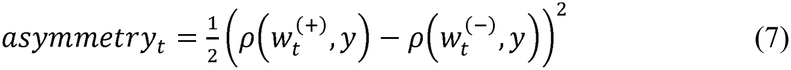

To calculate adjusted fit, we add asymmetry to fit and multiply this sum by *K*, the number of predictive variables included in the prediction, as shown by equation 8. Adjusted fit recognizes that in the context of evaluating subsets of variables, we are more likely to observe a spurious relationship between prediction weights and outcomes based on just one or a few variables than we are based on a collection of many variables. This comment warrants elaboration. We recognize that in the context of selecting all the variables that inform an entire prediction routine, including more variables increases the possibility of obtaining a spurious result. However, we must multiply fit by K to address an inherent property of fit. If we do not multiply fit by K, introducing an irrelevant variable such as a pure noise variable mechanically reduces fit. Multiplication by K allows for the inclusion of additional variables without penalizing them irrespective of their predictive value. This multiplication by K puts fit in the same units as R-squared, as can also be seen from our equivalence proof in the appendix.

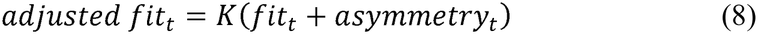

RBP forms a prediction as a weighted average of previous case outcomes in which the weights are based on the relevance of the previous cases. As we noted earlier, if we use the full sample of previous cases, RBP gives the same prediction as linear regression analysis.

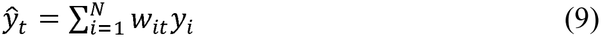

In equation 9, *ŷ*_*t*_ is the prediction for the current case, *y*_*i*_ is a previous case, *N* is the number of cases in the full sample, and *w*_*it*_ is the relevance-based weight as defined by equation 10. Equation 10 shows that a case’s relevance is used to tilt the case’s weight away from its average weight in the sample, recalling that relevance is centered on zero.N

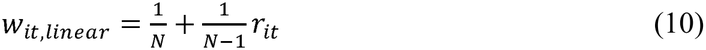

Fit allows us to produce a more reliable prediction by censoring cases that are less relevant than a chosen threshold. Equations 11 through 13 show how we weight previous cases when we censor some of them.

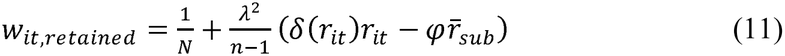

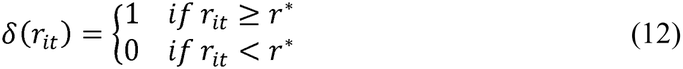

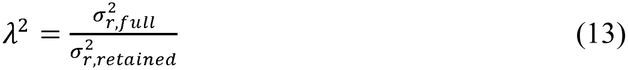

In equations 11 through 13, 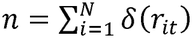 is the number of cases that are fully retained, which is to say, those cases whose relevance is equal to or exceeds the relevance threshold, *φ* = *n*/*N* is the fraction of cases in the retained sample, and 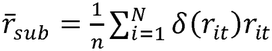 is the average relevance value of the cases in the retained sample. It is important to note that *w*_*it*_,*retained* depends crucially on the prediction circumstances *x*_*t*_. Relevance is reassessed for each prediction circumstance which further affects the identification of the retained subsample and introduces nonlinear conditional dependence of the prediction *ŷ*_*t*_ on the prediction circumstances *x*_*t*_. The scaling factor *λ*^2^, which is measured as the ratio of the variances of relevance within the full and retained samples, compensates for a bias that would otherwise result from relying on a small subsample of highly relevant cases. In the case of linear regression *n* = *N* and *λ*^2^ = 1. Lastly, note that the regression weights always sum to 1.^18^

### Grid Prediction

Thus far, we have implicitly assumed that we always use the same set of predictive variables and only consider different subsamples of relevant cases. But it is unwise to assume that a fixed set of predictive variables will be equally effective across all subsamples of cases. And it is equally unwise to assume a fixed subsample of cases will yield the best results irrespective of the chosen predictive variables. Instead, we must recognize that the choice of cases and the choice of predictive variables are codependent on the specific circumstances of each prediction task.

This codependence is best illustrated by a grid in which the columns represent different combinations of predictive variables, and the rows represent subsamples of cases determined by different relevance thresholds. Each cell contains a prediction and its reliability weight. Grid prediction forms a composite prediction as a reliability-weighted average of the predictions from all possible calibrations. Equation 14 defines reliability weights, *ψ*_*θ*_, as the adjusted fit for a parameter calibration, *θ*, divided by the sum of all adjusted fits across all parameter calibrations.

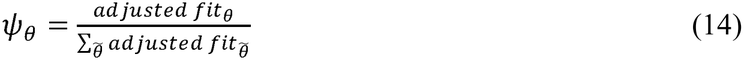

We form a composite prediction by taking a weighted average of a representative set of cells in which the weights are based on the adjusted fits of the predictions, as shown by equation 15. By forming a composite prediction, we guard against data errors or overreliance on potentially tenuous information.

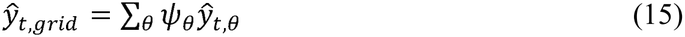

An exhaustive search of the grid could be computationally infeasible, which is why we sample the grid to form a composite prediction that is both robust and efficient to compute. Alternatively, we could search the grid and use the cell with the highest fit to form our prediction. This approach, however, concentrates the prediction on a single calibration which may include an important data error, or which may give a very different prediction from another cell whose fit is nearly as high. In any event, by diversifying the composite prediction across a reasonable number of cells, we are less vulnerable to data errors or spurious relationships because many specific problems will be confined to a limited number of grid cells.

It is important to note that the prediction grid allows us to filter observations based on both relevance, which contains similarity and informativeness, as wells as similarity by itself. Although information theory shows that unusual observations contain more information than common observations, it is sometimes the case that extrapolation from unusual observations is not warranted. In such instances it may be preferable to filter by similarity. Fortunately, we need not worry which filter to apply. We rely on fit to make this determination. That said, we always form the prediction weights based on relevance.

We also wish to note that the relevance thresholds, variable combinations, and reliability weights are determined independently of prediction outcomes. RBP is principle based as opposed to a model whose calibration requires empirical validation.

Given that a representative prediction grid across many variables is too large to show, we provide a visual representation of a hypothetical prediction grid in Table 1. Each cell represents a calibration *θ*; that is, a unique combination of predictive variables and cases. The first values shown in the cells are the calibration-specific predictions *ŷ*_*t*_ for a given prediction task *t*. The second values are the weights *ψ*_*θ*_ we apply to the calibration-specific predictions to form the composite prediction. In this illustration the grid includes censoring thresholds at increments of 0.1. In our actual analysis that follows, we construct a grid with censoring thresholds of 0, 0.2, 0.5, 0.8, 0.9, and 0.99 to allow for a concise collection of filtered subsets that nevertheless span a wide range in terms of selected subsample size.

**Table 1.**
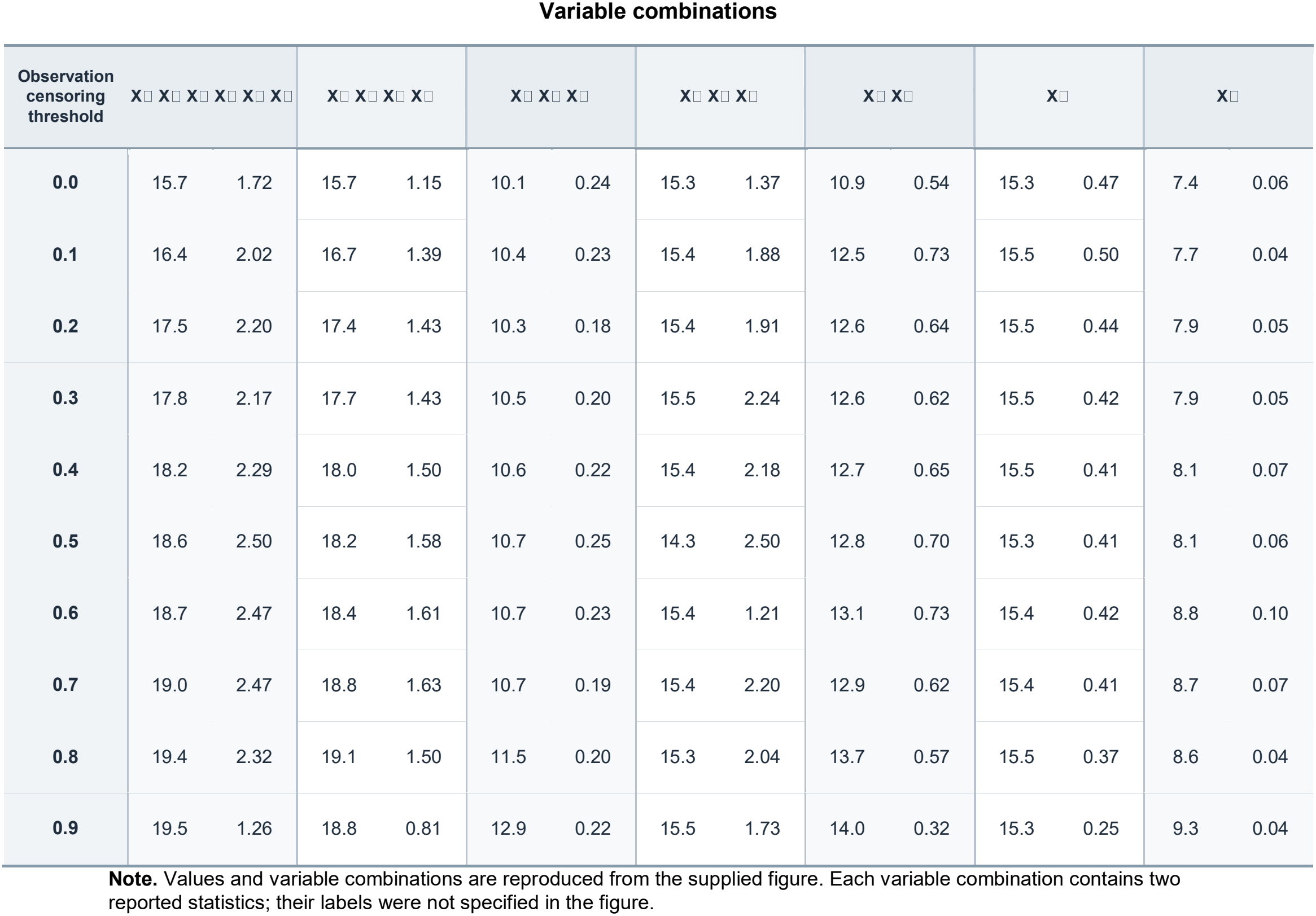
Prediction Grid: A Visual Representation of a Hypothetical Prediction Grid.

| Observation censoring threshold | X□ X□ X□ X□ X□ X□ |  | X□ X□ X□ X□ |  | X□ X□ X□ |  | X□ X□ X□ |  | X□ X□ |  | X□ |  | X□ |  |
|---|---|---|---|---|---|---|---|---|---|---|---|---|---|---|
| **0.0** | 15.7 | 1.72 | 15.7 | 1.15 | 10.1 | 0.24 | 15.3 | 1.37 | 10.9 | 0.54 | 15.3 | 0.47 | 7.4 | 0.06 |
| **0.1** | 16.4 | 2.02 | 16.7 | 1.39 | 10.4 | 0.23 | 15.4 | 1.88 | 12.5 | 0.73 | 15.5 | 0.50 | 7.7 | 0.04 |
| **0.2** | 17.5 | 2.20 | 17.4 | 1.43 | 10.3 | 0.18 | 15.4 | 1.91 | 12.6 | 0.64 | 15.5 | 0.44 | 7.9 | 0.05 |
| **0.3** | 17.8 | 2.17 | 17.7 | 1.43 | 10.5 | 0.20 | 15.5 | 2.24 | 12.6 | 0.62 | 15.5 | 0.42 | 7.9 | 0.05 |
| **0.4** | 18.2 | 2.29 | 18.0 | 1.50 | 10.6 | 0.22 | 15.4 | 2.18 | 12.7 | 0.65 | 15.5 | 0.41 | 8.1 | 0.07 |
| **0.5** | 18.6 | 2.50 | 18.2 | 1.58 | 10.7 | 0.25 | 14.3 | 2.50 | 12.8 | 0.70 | 15.3 | 0.41 | 8.1 | 0.06 |
| **0.6** | 18.7 | 2.47 | 18.4 | 1.61 | 10.7 | 0.23 | 15.4 | 1.21 | 13.1 | 0.73 | 15.4 | 0.42 | 8.8 | 0.10 |
| **0.7** | 19.0 | 2.47 | 18.8 | 1.63 | 10.7 | 0.19 | 15.4 | 2.20 | 12.9 | 0.62 | 15.4 | 0.41 | 8.7 | 0.07 |
| **0.8** | 19.4 | 2.32 | 19.1 | 1.50 | 11.5 | 0.20 | 15.3 | 2.04 | 13.7 | 0.57 | 15.5 | 0.37 | 8.6 | 0.04 |
| **0.9** | 19.5 | 1.26 | 18.8 | 0.81 | 12.9 | 0.22 | 15.5 | 1.73 | 14.0 | 0.32 | 15.3 | 0.25 | 9.3 | 0.04 |

**Note.** Values and variable combinations are reproduced from the supplied figure. Each variable combination contains two reported statistics; their labels were not specified in the figure.

Note that each cell’s prediction is a linear function of cases, and the grid prediction is a linear function of each cell’s prediction. Therefore, we can express the grid prediction in terms of composite weights applied to each case, as shown by Equation 16. Composite weights are important because they preserve the transparency of each case’s contribution to the current prediction task, and they allow us to calculate fit from composite weights as a final gauge of the grid prediction’s reliability.

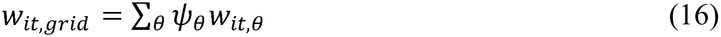

In equation 17, we compute the fit of the composite prediction as the squared correlation of the composite prediction weights with the outcomes, and we multiply by weighted average of the number of variables in each cell, *K*_*θ*_, which represents the effective number of variables used in the composite prediction, for the same reason that we multiply by *K* to compute adjusted fit, as discussed earlier.

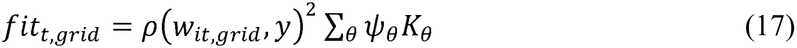

### Relevance-Based Importance

The prediction grid yields a comprehensive measure of how each variable contributes to a prediction’s reliability. Specifically, the Impact on Fit (IOF) of variable *k* is given by the average adjusted fit, *F*_*tθ*_, for grid cells that include variable *k* (Δ_*k*_(*θ*) = 1) minus the average adjusted fit for the remaining cells (Δ_*k*_(*θ*) = 0), as shown by equation 18.

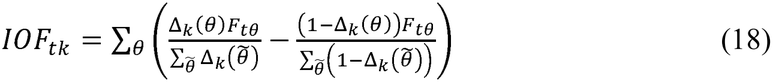

The term 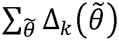 in the denominator counts the number of cells that include variable *k*. For a grid that includes every variable combination, this number is nearly equal to the number of cells that do not include variable *k*, but the counts are not identical unless we include a column in the grid for predictions that do not use any of the *X* variables (for which adjusted fit is always zero). Thus, we divide by the number of cells that include variable *k* regardless of whether a given cell contains *k* or not.

The value of IOF may be positive, zero, or negative. A positive value indicates that a variable adds value to the reliability of a prediction, and the higher the value the more substantial its contribution. A zero, or near-zero, value indicates that a variable contributes benign noise to a prediction. A negative value indicates that a variable contributes harmfully.

IOF has several advantages over alternative measures of variable importance. Linear regression relies on t-statistics and their corresponding p-values, which only measure a variable’s marginal importance. IOF, by contrast, captures a variable’s total importance. IOF also captures conditional relationships which t-statistics fail to address. And unlike the Shapley value, which is the accepted standard for assessing variable importance in machine learning models, IOF accounts for the reliability of individual predictions.

We can use the same approach to measure how each variable contributes to the value of a prediction. This measure is called Impact on Prediction (IOP), and it is given by the average prediction from the cells including variable *k* minus the average prediction from the remaining cells, as shown by equation 19.

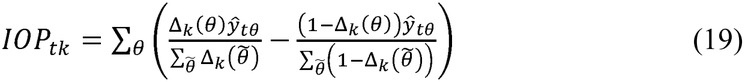

It should be noted that although equation 19 gives a useful indication of the contribution of each variable, it does not constitute an exact decomposition of the grid prediction value. To calculate an exact decomposition would require forming new composite grid predictions for every subset of variables.

### Distribution of Information

As we previously mentioned, fit quantifies the strength of the patterns in a dataset that is used to form a prediction, which we can think of as the signal that supports the prediction. We now show how to represent the noise that surrounds the signal, which we refer to as the distribution of information. Suppose we use equation 11 to retain only a single case and form a prediction from equation 9. To enable this calculation, we first rewrite the combined form of these two equations in a form that removes the explicit division by *n* − 1 and expresses the prediction’s deviation from the full sample average as a weighted average of the observed deviations from average. In equation 20, we express 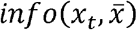 as *info*( *x*_*t*_) for concision.

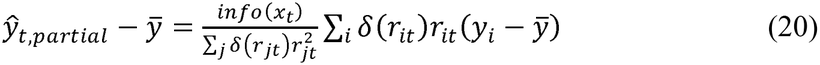

We retain just a single case to arrive at the prediction in equation 21, which we call a solo prediction because it relies on deviations from average for just one case.

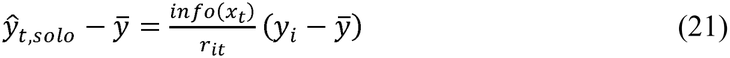

The solo prediction for case *i* scales the deviation 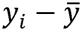 by the informativeness of the prediction circumstances and divides by the relevance of the case to the prediction circumstances. We can think of the multiplication by informativeness as scaling the observed deviation to predict larger effects for more extreme circumstances. The division by relevance accomplishes two goals. First, it can flip the direction of the deviation if the case is opposite to the circumstances. Second, it further scales the deviation by identifying the extent to which the case is aligned to current circumstances. For example, if the case’s predictive variables exactly equal the circumstances, the ratio is one and the prediction is the deviation in outcomes. To the extent the observation does not align with the circumstances there is a smaller signal in the case, so we boost it further to align with the expected size of the future outcome.

A solo prediction by itself would have exceptionally low fit because it relies on such a narrow sample. It would be unreliable. However, we make use of solo distributions by reformulating the prediction as a weighted average of them. We solve for the weights that generate the prediction, as given by equation 22. Thus, the weight placed on the solo prediction of a retained case is equal to its squared relevance as a fraction of the squared relevance of all the other retained cases. These weights sum to one, they are all nonnegative, and the weights of the censored cases in this context are all zero.

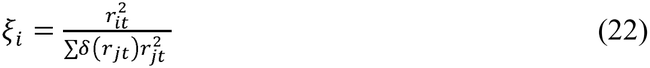

By embedding the scaling and directionality of each extrapolation and expressing deviations from average, solo predictions convert a weighted average that potentially contains negative weights to a new weighted average that does not. We can interpret these positive weights that sum to one as frequencies and compile an empirical distribution of the information that comprises a prediction. To apply this method to the prediction grid, we pool the weights and solo predictions across cells, weighted by their adjusted fit, and compute a composite frequency distribution.

Before proceeding to our illustration of RBP, we would like to further motivate its application by reviewing its theoretical justification and by comparing its transparency to several alternative prediction methods.^19^

As we have shown, RBP forms a prediction as a weighted average of observed outcomes with weights determined by a precisely defined statistic called relevance. The theoretical motivation of relevance draws on information theory, the Central Limit Theorem, and the Mahalanobis distance. Information theory holds that the information contained in an observation is the negative logarithm of its probability, implying that unusual observations contain more information than common observations. The Central Limit Theorem provides motivation for using a multivariate normal distribution to characterize data for the purpose of initially determining relevance. The normal distribution provides a natural prior because it is the maximum entropy distribution for a given mean and covariance matrix. It therefore represents the broadest distribution of uncertainty consistent with these constraints without imposing additional assumptions. And setting aside the constants, the negative logarithm of the multivariate normal density equals one-half the squared Mahalanobis distance. This relationship provides the theoretical justification for incorporating informativeness into relevance. That relevance gives a principled way to form prediction weights is further justified by the following mathematical equivalences. When RBP is implemented using the full sample of observations, it produces the same prediction as linear regression. And under the Gauss-Markov assumptions, ordinary least squares provides the best unbiased estimator. And as we have mentioned, fit, which depends on relevance and guides the selection of observations and variables, is an exact decomposition of R-squared in linear regression. Having reviewed the theoretical foundations of RBP, we now compare it to several alternative prediction methods to further clarify RBP’s distinctive features.

### RBP versus Neural Networks

A neural network uses large numbers of parameters to model nonlinear relationships through sequences of linear combinations and nonlinear transformations. They are highly flexible and capable of identifying complex relationships in large datasets. However, their specification and structure are determined empirically rather than deduced from theory or first principles. As a result, they are susceptible both to overfitting spurious relationships and to underfitting important but rare relationships.

RBP is a principle-based prediction routine as opposed to an empirically determined model. RBP forms new prediction weights for each task whereas a neural network uses the same parameters to address all prediction tasks. Therefore, RBP only needs to address the conditionalities relevant to a single prediction circumstance. This feature gives RBP the power to capture the same nonlinear and conditional relationships as a neural network in most cases.

Transparency is RBP’s most distinctive feature compared to neural networks. Neural networks are often characterized as black boxes because they provide little insight into how observations or variables influence predictions. RBP, by contrast, explicitly reveals how each observation contributes to each prediction and how each variable contributes to the value and reliability of each prediction.

Also, neural networks do not give advance notice of a specific prediction’s reliability. This information is known only after we observe the outcomes. By comparison, RBP uses fit to assess each prediction’s reliability beforehand, and it also reveals in advance the distribution of solo predictions which shows whether the underlying observations provide mutually reinforcing or contradictory information.

### RBP versus Gaussian Kernels and k-Nearest Neighbors

A Gaussian kernel forms predictions as weighted averages of nearby observations, in which the weights decline smoothly according to a Gaussian distribution the further they are from the prediction circumstances. The weights that are used to form a prediction depend on the Euclidean distance and the width of the Gaussian distribution.

Although RBP shares the intuition that similar observations should matter more than dissimilar observations, it differs in several important respects. First, RBP measures similarity using the Mahalanobis distance rather than the Euclidean distance, allowing it to account for covariation among variables. Second, RBP incorporates informativeness in addition to similarity. Third, RBP adapts the relevance thresholds to each prediction task, using fit and the prediction grid for each specific prediction task, rather than applying one fixed weighting scheme to all prediction circumstances.

Gaussian kernels provide some transparency because they reveal the relative influence of observations. However, they do not evaluate reliability or variable importance, and they rely on a single calibration for all predictions, such as a fixed width of the Gaussian distribution. RBP’s grid prediction framework allows both observation weights and variable combinations to vary across prediction tasks, which improve flexibility and interpretability.

The related algorithm k-nearest neighbors predicts outcomes by averaging a fixed number of nearby observations and weighting them equally. Relative to both Gaussian kernels and k-nearest neighbors, RBP is both more rigorous and transparent.

### RBP versus XGBoost

XGBoost is a gradient boosting algorithm that builds an ensemble of decision trees sequentially. Each new tree attempts to correct the residual errors made by previous trees, and the final prediction is the sum of the trees’ outputs. XGBoost is highly flexible and effective at capturing nonlinear relationships. XGBoost relies primarily on empirical calibration rather than theoretical structure. Its parameter values are optimized according to average performance across training data, thereby rendering it vulnerable to spurious relationships when future conditions differ from those represented in the training sample.

RBP addresses complex conditional relationships differently. Instead of fitting a global model to all tasks, it evaluates a vast number of combinations of observations and variables for each specific prediction. This task-specific approach allows RBP to adapt to varying prediction circumstances while remaining transparent. XGBoost can quantify how predictive variables influence individual predictions using methods such as SHAP, but it does not inherently reveal how each training observation contributes to a prediction or how each variable affects its reliability. Moreover, conventional XGBoost predictions do not inherently provide a prediction-specific measure of reliability. RBP, by contrast, explicitly reveals how the observations and variables influence each prediction, and it gives an estimate of each prediction’s reliability based on the fit associated with the predictions from a vast number of combinations of observations and predictive variables.

## Application of RBP to Predict Opioid Outcomes

We now illustrate RBP by predicting impact scores for patients who were treated with opioids. We first describe our data and experiment set up. Then we present our results. Again, we wish to stress that RBP can be applied to any data-driven prediction task.

### Data Source and Availability

Data were provided by Celéri Health, Inc. to Cambridge Prediction Analytics under a data use agreement (DUA). The analytic extract was limited to opioid initiation episodes and comprised 119,966 follow-up assessments across 16,920 episodes in 13,641 patients from 36 states. Each episode required a baseline PROMIS-29 within 30 days before the opioid start date; follow-up assessments were serial, with a median of four per episode. Records used include gender, age, PROMIS-29 domain T-scores, global pain rating, and impact score, with all timing expressed as day intervals relative to the opioid start date.

For the present analysis, inclusion criteria were limited to those who had initial baseline values and follow-up observations were restricted to those occurring more than 180 days and less than 260 days after the opioid start date. Observations at exactly 180 or 260 days were excluded. We selected the 180–260 day window to obtain a reasonably consistent measure of treatment outcomes approximately six to eight months after treatment. Assessments conducted earlier may not fully capture the treatment effect, while assessments conducted substantially later may be more susceptible to intervening changes in patient health, additional treatments, or other confounding factors. Restricting the assessment window also improves comparability across patients by limiting variation in follow-up duration. Patients of age 90 and older were excluded. This filtering yielded 11,543 observations. Since some patients had more than one qualifying follow-up observation, the data set was collapsed to the episode level by retaining the earliest qualifying follow-up observation for each patient. This filtering yielded 7,609 observations. Lastly, we removed any observations that represent a second or subsequent treatment for each patient. The final analytic cohort included 5,851 episodes in which each patient is included only once. Episodes were excluded if they did not meet the above criteria. The data are proprietary to Celéri Health, Inc. and were accessed under the above DUA that does not permit redistribution of patient-level records. Celéri Health had no role in the study design, analysis, interpretation, or decision to publish, and does not endorse the methodology or conclusions reported here.

### Experiment Setup and Predictive Variables

Using RBP, we predict the impact score that occurs approximately six months following opioid treatment for each of 5,851 cases. Specifically, we identify every case for whom at least one impact score assessment is available between 180 and 260 days following treatment, and we record the earliest assessment within that time window as the case’s outcome of interest. We use a “leave one out” method where each case is predicted based on the observed outcomes from the other 5,850 cases. As mentioned previously, each patient only has one case in the data sample, so the leave one out test ensures that no patient’s own prior outcomes could ever be used to predict his or her outcome. There is no time dimension to our analysis. Lower values indicate lower impact scores (more desirable outcomes). We include the baseline impact score prior to treatment as a predictive variable (essentially, a control). We used 11 predictive variables to form our predictions of impact score (Table 2). The values of predictive variables were available as of the time of first opioid treatment initiation, collected within the prior 30 days as mentioned above, and the outcomes reflect measurements made at the pertinent follow up visit. This entire setup is refreshed for each new prediction task.

**Table 2.**
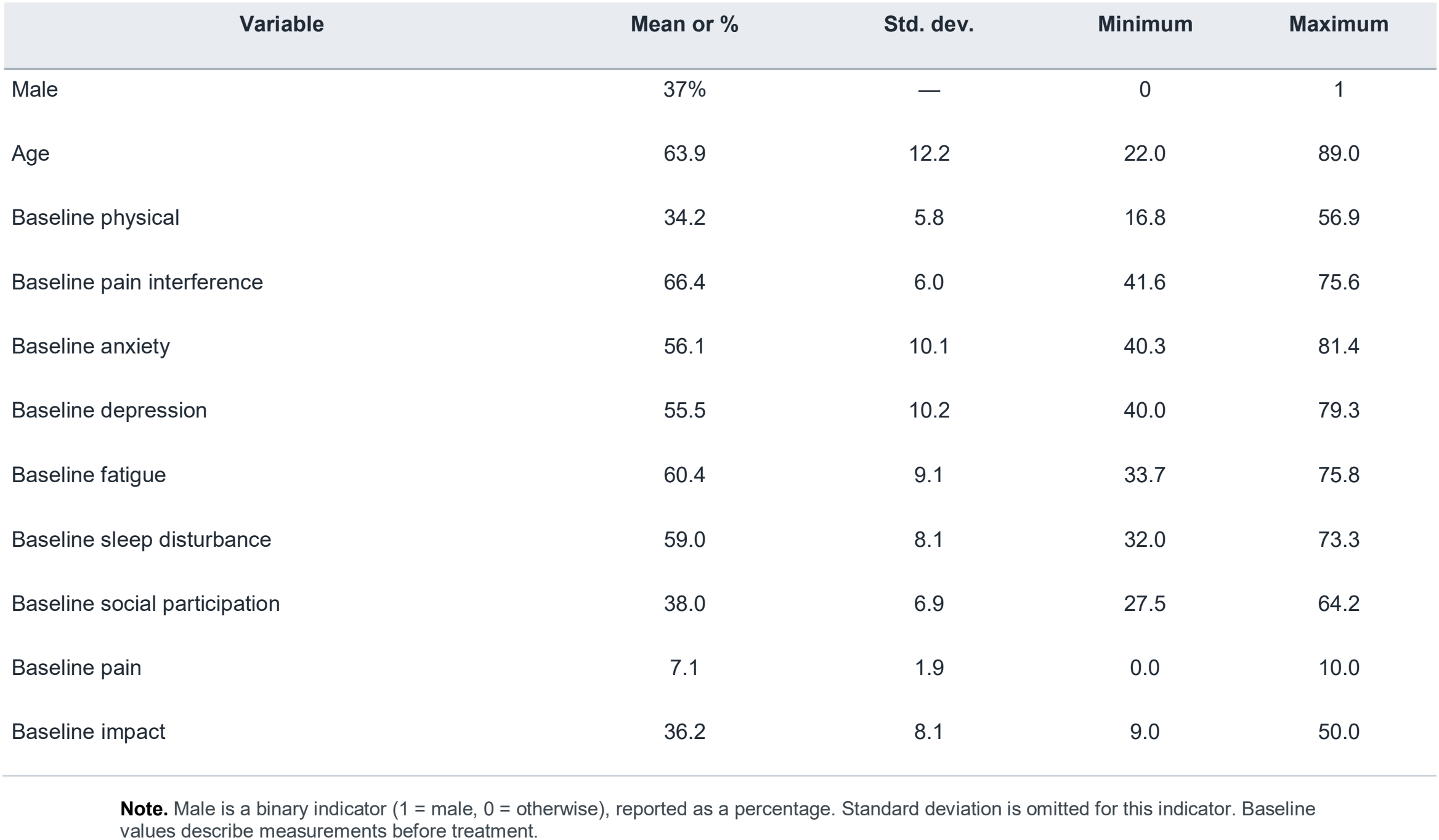
Descriptive Statistics of the Study Sample (N = 5,851)

## Results

### Predictions and Reliability

Our first set of results shows the predictions of impact score for each case expressed as cross-sectional percentile ranks on the vertical axis along with the reliability RBP assigns to each prediction based on fit (equations 6 – 8) and presented as cross-sectional percentile ranks on the horizontal axis (Figure 1). It reveals that for a given level of prediction, there can be a wider dispersion of reliability because there are more robust patterns to support some predictions than others. It is interesting to note that as the level of reliability decreases, the predictions converge toward the average impact score, which is what we should expect to observe. If we lack confidence in our ability to predict an outcome, we should default to the average value. It is also important to note that the information given by the horizontal axis is unavailable from a regression model, for example, which assumes that all predictions are equally reliable.

**Figure 1:**
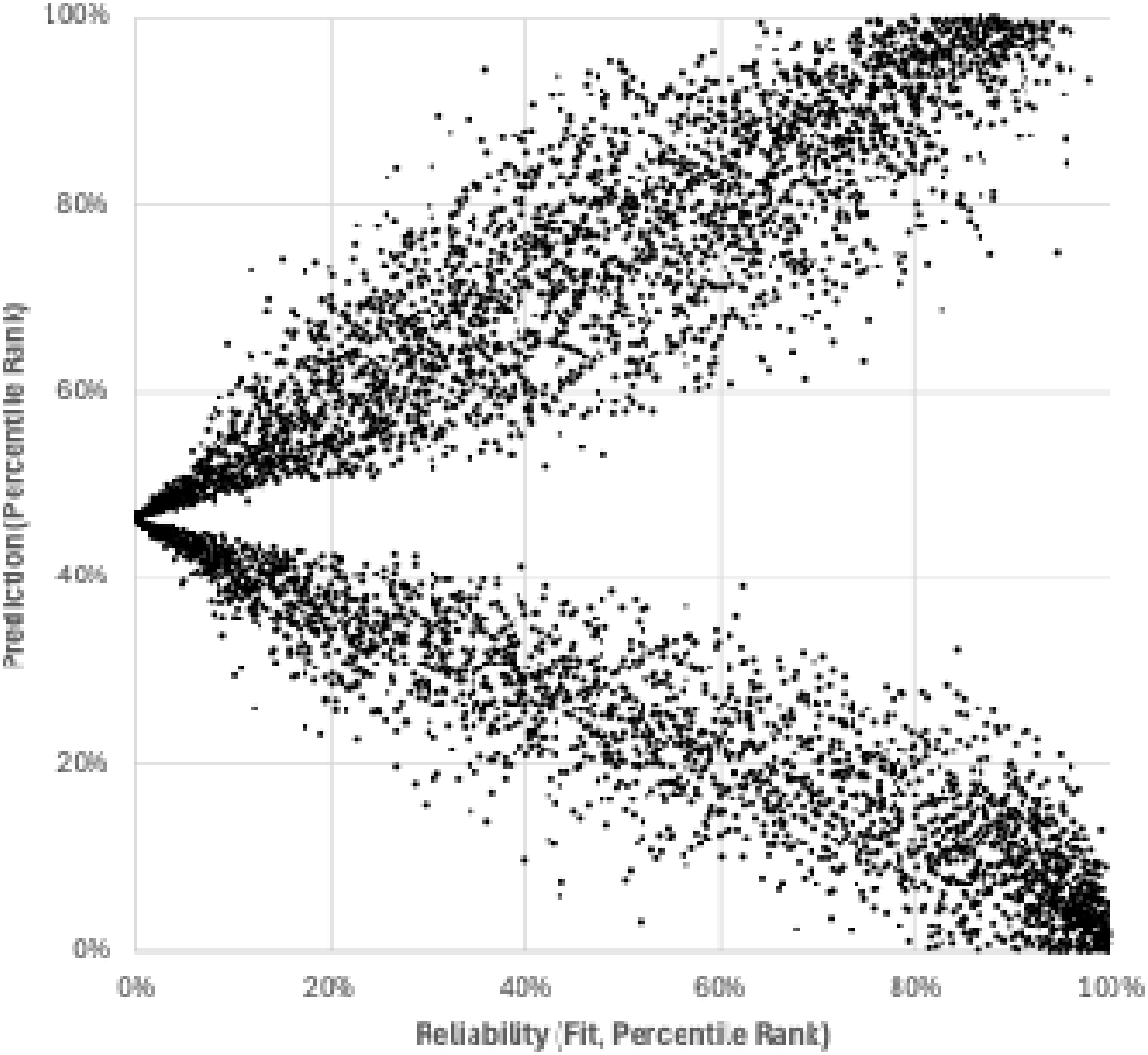
Scatter Plot of Impact Score Predictions and Reliability (Fit) for 5,851 Cases.

### Prediction Efficacy

We give evidence of prediction efficacy (Table 3). We observe the realized outcomes of impact score for the half of the sample we predicted would report above average impact scores and for the other half of the sample we predicted would report below average impact scores for the full set of predictions as well as for subsamples of higher and lower fit known in advance of the predictions.

**Table 3.** Realized Outcomes by Predicted Outcome and Prediction-Specific Fit.

|  | Linear | RBP | RBP 50% |  | RBP 25% |  | RBP 10% |  |
|---|---|---|---|---|---|---|---|---|
|  | Regression | All | Highest fit | Lowest fit | Highest fit | Lowest fit | Highest fit | Lowest fit |
| **Average outcome (full-sample mean = 35.0)** |  |  |  |  |  |  |  |  |
| Top half | 39.8 | 39.5 | 41.3 | 37.7 | 42.3 | 36.8 | 42.6 | 36.3 |
| Bottom half | 30.6 | 30.5 | 27.3 | 33.7 | 25.1 | 34.8 | 22.8 | 35.4 |
| **Spread (top − bottom)** | **9.2** | **9.0** | **14.0** | **4.0** | **17.2** | **2.0** | **19.8** | **0.9** |
| **95% confidence interval for average outcome** |  |  |  |  |  |  |  |  |
| Top half | [39.5, 40.0] | [39.3, 39.8] | [41.0, 41.6] | [37.4, 38.1] | [41.8, 42.8] | [36.3, 37.2] | [41.8, 43.4] | [35.5, 37.0] |
| Bottom half | [30.3, 30.8] | [30.2, 30.8] | [26.9, 27.7] | [33.4, 34.0] | [24.6, 25.6] | [34.3, 35.2] | [22.0, 23.6] | [34.6, 36.0] |
| **Spread (top − bottom)** | [8.9, 9.6] | [8.6, 9.4] | [13.5, 14.5] | [3.6, 4.5] | [16.4, 17.9] | [1.3, 2.7] | [18.7, 20.9] | [−0.1, 1.9] |
| **Root mean squared error (RMSE)** |  |  |  |  |  |  |  |  |
| Top half | 4.8 | 4.8 | 4.8 | 4.7 | 4.9 | 4.8 | 5.1 | 4.9 |
| Bottom half | 4.9 | 4.9 | 4.8 | 5.0 | 4.9 | 4.9 | 4.9 | 4.9 |
| **Spread (top − bottom)** | −0.1 | −0.1 | 0.0 | −0.2 | 0.0 | −0.1 | 0.2 | 0.0 |
| **Sample sizes** |  |  |  |  |  |  |  |  |
| Top half | 2,825 | 2,926 | 1,463 | 1,463 | 732 | 732 | 293 | 293 |
| Bottom half | 3,026 | 2,925 | 1,462 | 1,463 | 731 | 732 | 293 | 293 |
| **Total** | 5,851 | 5,851 | 2,925 | 2,926 | 1,463 | 1,464 | 586 | 586 |

**Note.** Top and bottom halves refer to observations with higher and lower predicted outcomes within each sample. RBP fit groups comprise the highest- or lowest-fit 50%, 25%, or 10% of cases. Spreads are differences between the top and bottom groups. The 95% confidence intervals are percentile intervals from 5,000 nonparametric bootstrap resamples of realized outcomes, drawn independently with replacement within each original group; for spreads, each resample subtracts the bottom-group mean from the top-group mean. Predictions and group assignments were held fixed. The intervals describe conditional sampling variability and do not include uncertainty from refitting the prediction methods or redefining the groups. Counts are numbers of predictions.

For the full sample of predictions, the realized spread in the average values of the impact scores between above average and below average predictions was 9.0, demonstrating that RBP effectively predicted impact scores. For those predictions known in advance to be the 50% most reliable the spread increased to 14.0, and for those predictions known ahead of time to be the 50% least reliable, it was only 4.1. When we consider the 25% most reliable predictions the spread in realizations between above average and below average predictions increased to 17.3, and for those predictions known to be the least reliable, it was only 1.7. The same pattern prevails when we consider the 10% most and least reliable predictions: the 10% most reliable predictions had a spread of 20.2 compared to 0.5 for the least reliable predictions.

Our choice of spread in outcomes as a measure of predictive efficacy deserves elaboration. We think it is important to evaluate prediction errors not only by their magnitude but also in the context of the information that supports the prediction. Consider, for example, two predictions: one which predicts a notably high score of 45 when the actual score is 40, and one which predicts a closer-to-average score of 35 when the actual score is 30. Although these errors contribute equally to RMSE, they may have different implications. In the first case, the error may have no consequence for a treatment decision whereas in the latter case, the error may guide the caregiver to change the treatment.

The differences shown in Table 3 in the spreads of the realizations for above and below average predictions across predictions with varying levels of reliability offer persuasive evidence of RBP’s ability to distinguish in advance more trustworthy predictions from those predictions that should be viewed with greater skepticism. Keep in mind that a regression model’s R-squared would assign the same reliability to the 10% least reliable predictions whose realizations had a spread of only 0.5 as it would to the 10% most reliable predictions whose realizations had a spread of 20.2.

The case-specific information about reliability given by RBP has a very important and practical benefit. RBP would guide a caregiver to be more cautious about giving risky treatment to a patient whose favorable prediction is known in advance to be highly unreliable than to a patient whose favorable prediction is known to be highly trustworthy. A regression model would guide a caregiver to treat both patients the same.

We again wish to emphasize that our focus in this paper is on transparency, though we acknowledge that predictive accuracy is an essential feature of any prediction method. Therefore, in addition to the information given by Table 3, we include references to two papers which compare the predictive efficacy of RBP to linear regression and machine learning models in other domains.^22,24^ In the next section we turn to our primary focus: how RBP provides explicit empirical support for individual predictions, thereby rendering them open to scrutiny.

### Prediction Formation

We now show how RBP gives visibility into the formation of a prediction which model-based prediction cannot do. The 10 most relevant cases for case A are illustrated which had a relatively low prediction but with high reliability and the 10 most relevant cases for case B which also had a low prediction but with lower reliability (Table 4). Thousands of cases were used to form each of these predictions, but we only show the 10 most relevant cases. Notice that while both cases have very similar predictions, their most relevant cases are completely different. The weights for case B are larger, indicating greater concentration on a relatively smaller sample of relevant cases, which may be one reason it has lower reliability.

**Table 4.** Prediction-Specific Results and Most Relevant Observations.

| Panel A. Prediction Results |  |  |  |  |  |  |
| Measure | Case A | Percentile | Case B | Percentile |  |  |
|---|---|---|---|---|---|---|
| **Prediction** | 27.14 | 10% | 28.20 | 13% |  |  |
| **Reliability** | 1.41 | 94% | 0.82 | 72% |  |  |
| **Actual outcome** | 27 | 17% | 26 | 14% |  |  |
| **Panel B. Ten Most Relevant Observations** |  |  |  |  |  |  |
| **Rank** | **Case A** |  |  | **Case B** |  |  |
|  | **Case ID** | **Weight** | **Impact score** | **Case ID** | **Weight** | **Impact score** |
| 1 | 1672 | 0.259% | 27 | 215 | 0.289% | 20 |
| 2 | 4510 | 0.231% | 15 | 4280 | 0.262% | 28 |
| 3 | 3684 | 0.221% | 15 | 5294 | 0.257% | 39 |
| 4 | 3816 | 0.220% | 20 | 3077 | 0.251% | 32 |
| 5 | 843 | 0.214% | 28 | 447 | 0.244% | 22 |
| 6 | 37 | 0.211% | 14 | 57 | 0.243% | 22 |
| 7 | 4792 | 0.208% | 23 | 2284 | 0.226% | 22 |
| 8 | 3021 | 0.200% | 30 | 51 | 0.218% | 27 |
| 9 | 1723 | 0.185% | 18 | 271 | 0.204% | 23 |
| 10 | 223 | 0.182% | 17 | 1959 | 0.204% | 26 |

**Note.** Panel A reports predicted and realized impact scores and their sample percentiles, together with prediction-specific reliability. Panel B lists the ten observations receiving the highest relevance weights for each prediction; weights are reported as percentages and do not represent all observations contributing to a prediction.

Next, we show how RBP gives visibility into the influence of each predictive variable on the reliability and magnitude of specific predictions (Figure 2). We selected two cases with similar impact scores but different levels of reliability to illustrate how RBP provides prediction-specific information about the importance of the observations used to form each prediction. The patient-specific contribution of the underlying observations is less observable in conventional model-based approaches.

**Figure 2:**
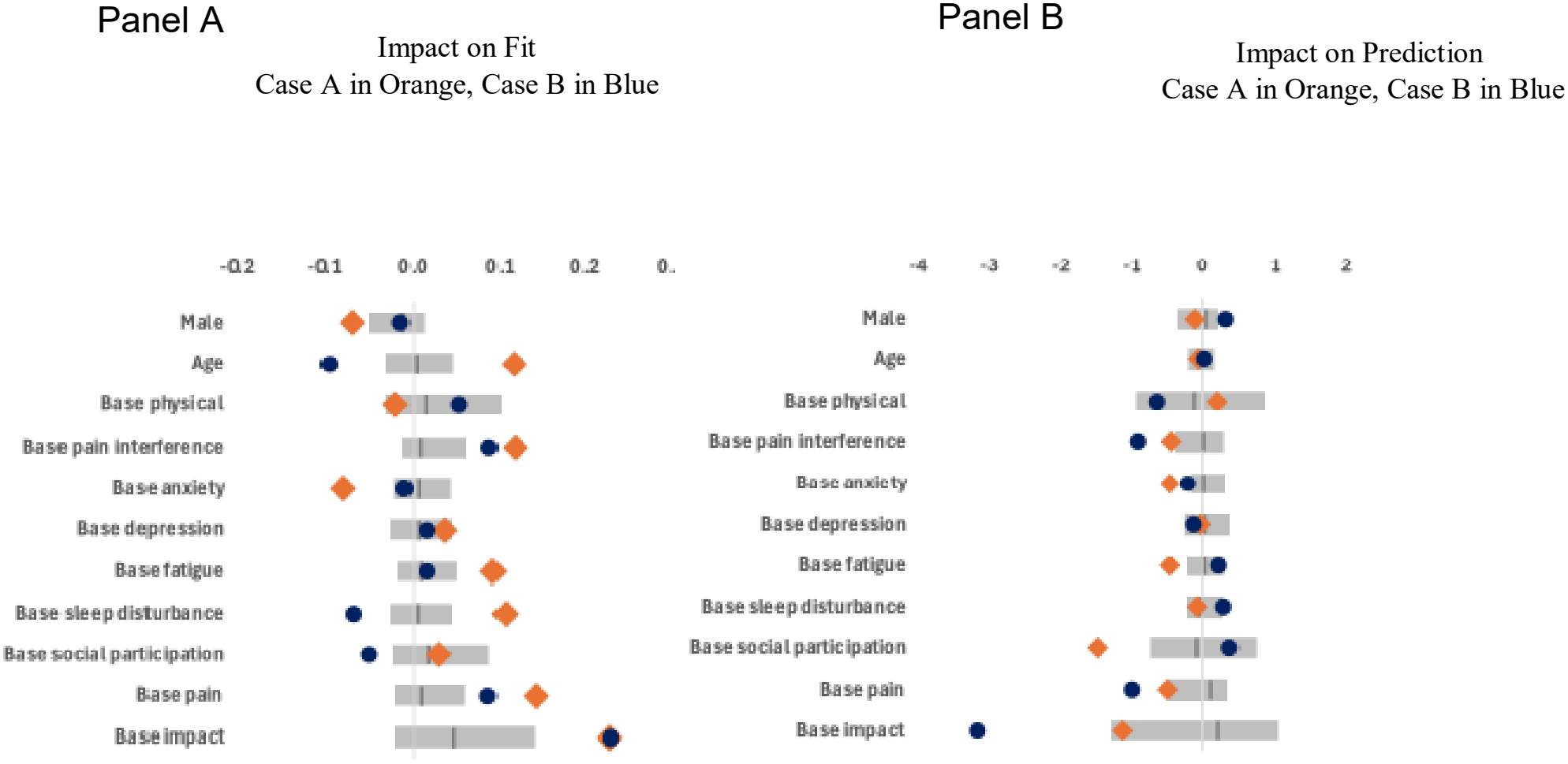
The Influence of Predictive Variables for Case A and Case B Relative to All Cases. A. Demonstration of how each variable contributes to the reliability of the predictions and B. how the variables impact the values of the predictions. The gray bars show the 20^th^ to 80^th^ percentile range (and median) across all predictions. The blue circles pertain to case B while the orange diamonds pertain to case A.

Figure 2A shows how each variable contributes to the reliability of the predictions, while Figure 2B shows how the variables impact the values of the predictions. A positive value indicates that including the variable in grid cells increases fit or the value of the prediction, respectively. If the value is zero it has no impact. If it is negative, including the variable reduces fit or the value of the prediction, respectively. The gray bars show the 20^th^ to 80^th^ percentile range (and median) across all predictions. The blue circles pertain to case B while the orange diamonds pertain to case A. These results highlight the importance of considering case-specific outcomes. Notice, for example, that age, base sleep disturbance, and base social participation decrease the reliability of the predictions for case B while they have the opposite effect for case A (Figure 2A). Also, notice that there is more dispersion, on balance, in how the variables impact the values of the predictions for case B compared to case A, which aligns with our earlier finding that the prediction for case B is less reliable than the prediction for case A (Figure 2B).

It is worth contrasting this case-specific information about the influence of predictive variables given by RBP with average-based information given by a regression model (Table 5). We therefore used linear regression to predict impact scores based on the same cases and predictive variables we used with RBP. Whereas regression coefficients and t-statistics describe relationships that are estimated across the sample, RBP provides additional information about how predictive variables influence the value and reliability of individual predictions. We highlight a few case-specific insights to illustrate this distinction. First, the t-statistic of −8.05 for base social participation conveys a strong negative relationship with outcomes after accounting for other variables, but it does not indicate whether base social participation improves the reliability of an individual prediction. RBP reveals that base social participation improves the fit of the prediction for case A and reduces the fit for case B. Similarly, the regression coefficient of −0.15 implies a static negative relationship between base social participation and the predicted outcome, holding other variables constant. By contrast, RBP shows that including this variable lowers the predicted impact score for case A but raises it for case B. These examples illustrate how RBP reveals prediction-specific relationships that are not directly captured by the global coefficients and t-statistics of a conventional linear regression.

**Table 5.**
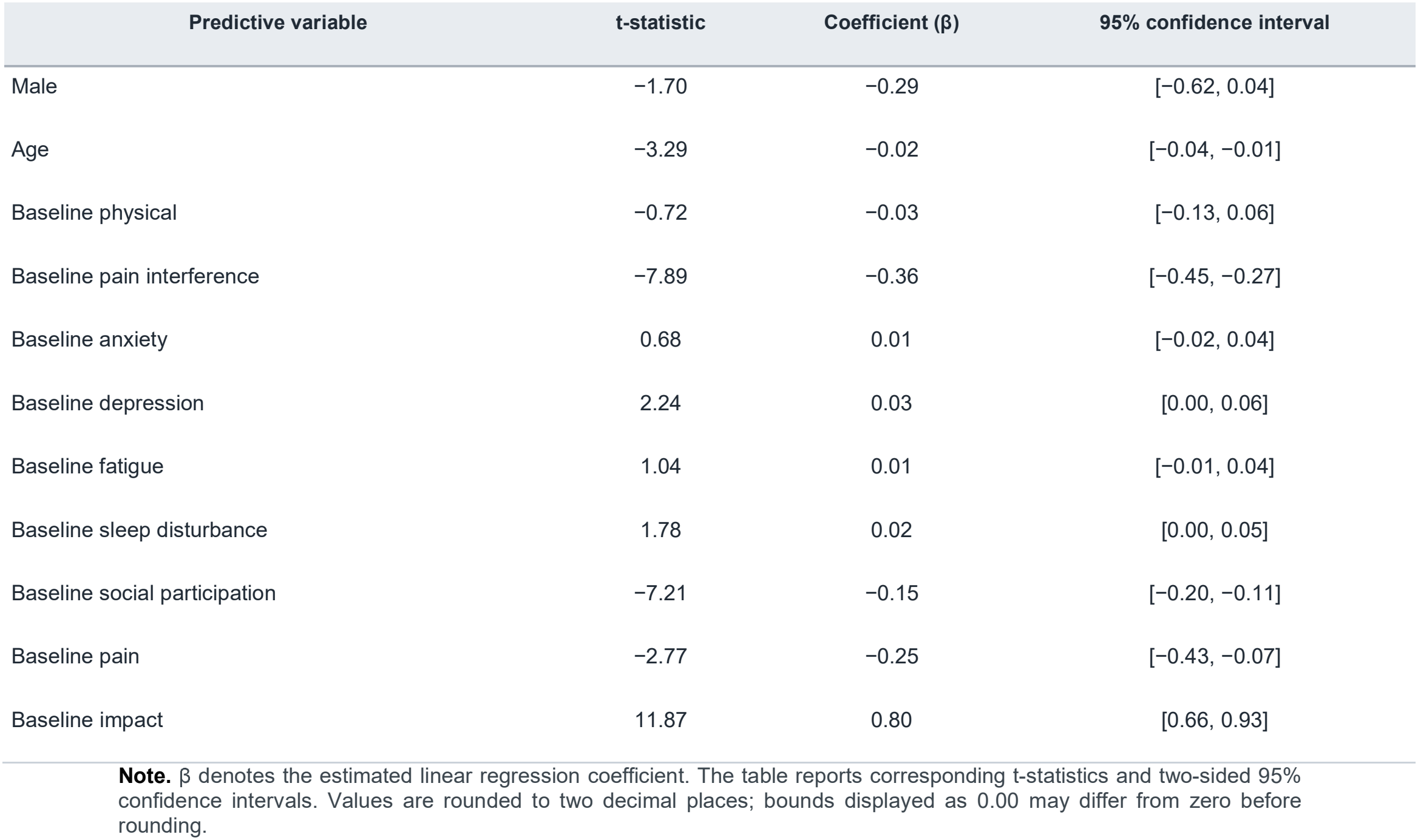
Linear Regression Results for the Full Sample.

### The Distribution of Information

We illustrated that RBP gives an indication of each prediction’s unique reliability as measured by fit in contrast to a regression model, for example, whose R-squared implicitly treats all predictions as equally reliable (Figure 1). As discussed previously, fit quantifies the strength of the patterns in the data used to form each prediction. Fit can be thought of as the strength of the signal that supports each prediction. Moreover, compelling evidence was provided of fit’s ability to distinguish more reliable predictions from less reliable predictions by reporting the gap in realized outcomes between cases predicted to have high impact scores versus those predicted to have low impact scores as a function of fit (Table 3). We now present a complementary measure of a prediction’s reliability by showing the distribution of the information that supports the predictions for case A and case B (Figure 3). These distributions represent deviations relative to the prediction value. The difference in the width of the bars is to enable comparison of the two distributions. They give a visualization of the noise that surrounds the signals given by fit for these two predictions. Notice that the solo predictions given by the observations for case B are more broadly dispersed than those for case A, indicating that there is less agreement among the observations for case B than there is for case A. This information reinforces the assessment that the prediction for case B is less trustworthy than the prediction for case A. Not only is the signal for case B weaker. It is also surrounded by more noise compared to case A as indicated by relatively higher probability bars for high and low outcomes.

**Figure 3:**
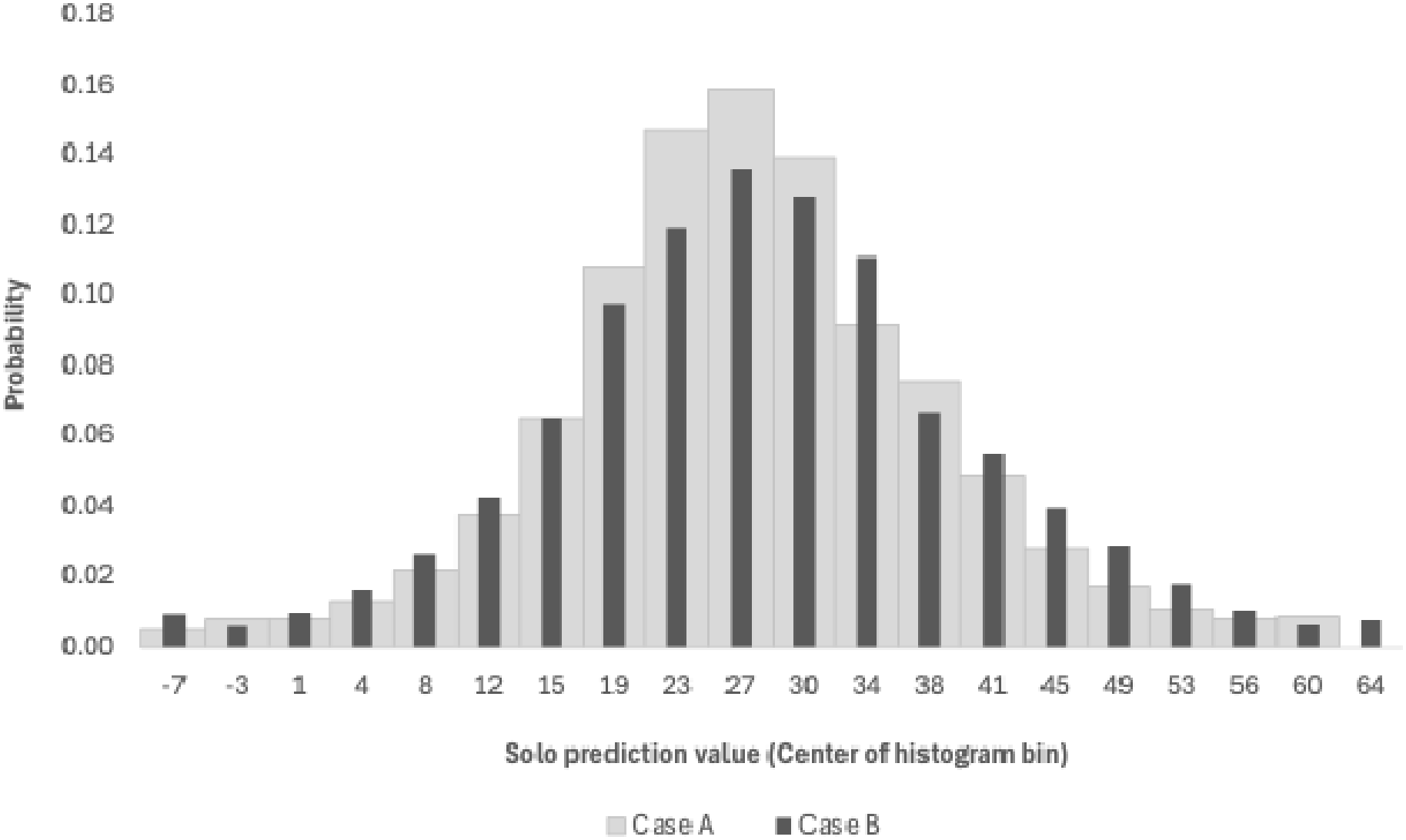
The Distribution of Information for Case A and Case B.

## Discussion

We introduced RBP which forms a prediction as a weighted average of observed outcomes in which the weights are based on a precise and theoretically justified statistic called relevance. We described the three key features of RBP: relevance, fit, and grid prediction. Relevance measures the importance of a case to a prediction and is composed of similarity and informativeness, which are both measured as MD. Cases that are like the current case but different from the average of all cases are more important to a prediction than those that are not. Fit quantifies the prevalence of useful patterns in a dataset and gives advance guidance about a prediction’s reliability. As such, fit determines the optimal blend of cases and predictive variables for each individual prediction task based on the unique circumstances of the prediction task. Grid prediction forms a composite prediction from many combinations of cases and predictive variables in a way that places more weight on combinations that are more reliable. Additionally, the prediction grid naturally yields comprehensive measures of variable importance. We also described how to measure the solo predictions given by individual cases, which enables us to view the distribution of information that supports each prediction which is the aggregation of the solo predictions. It is intuitive to think of fit as a measure of the strength of the signal that supports a prediction and the distribution of information as a visualization of the noise that surrounds the signal given by fit.

In an illustrative application, we demonstrated that RBP generates case-specific information about the reliability and formation of predictions of the impact score for patients treated with opioids that is unobservable from linear regression models. Specifically, RBP reveals the precise effect of each case on each prediction and the importance of each predictive variable to each prediction. Our analysis demonstrated that the influence of cases and predictive variables is highly prediction-specific, thereby highlighting that RBP offers valuable incremental insights to the information typically given by models.

The efficacy of medical treatment is often limited by reliance on model-driven summary information that ignores important case-specific information. For example, a regression model yields no information about the impact of specific cases on a prediction’s value beyond revealing that, on average, all the cases contribute equally to the value of the prediction, which is fruitless. Additionally, a regression model’s β coefficients assume that each variable’s impact on the value of a prediction is the same across all cases, and its t-statistics assume that each variable’s contribution to a prediction’s reliability also is the same across all cases. Finally, a regression model’s R-squared implicitly assumes that all predictions rendered by the model are equally reliable.

Since RBP is model-free and prediction-specific, it gives distinctive transparency into the formation and reliability of each prediction. For example, it reveals explicitly how each case informs each specific prediction. It shows how each predictive variable influences the reliability and value of each specific prediction. Most importantly, it assesses the reliability of each specific prediction before the prediction is rendered, thereby enabling clinicians to interpret and act on uncertain predictions with appropriate caution. Moreover, RBP has several additional advantages compared to model-based approaches to prediction. RBP is a theoretically grounded framework supported by information theory, the Central Limit Theorem, the MD, and important mathematical convergences. RBP is less prone to overfitting small samples in three ways. Fit optimally balances the stronger patterns that come from using a focused sample of more relevant cases with the additional noise that comes from shrinking the sample. Because RBP proceeds one task at a time, it needs only enough data to address the complexities associated with a specific task. And because RBP explicitly reveals how each case contributes to each prediction, it reveals how reliant a prediction is on one or just a few cases. Finally, even though it was not needed in this analysis because the data did not contain any missing values, when RBP is confronted with missing data it preserves information that a model must exclude or attempt to replicate. This occurs because data that is missing in one cell in the prediction grid will most likely be recovered in some of the other cells of the grid. Moreover, RBP explicitly accounts for the relative importance of missing data in forming a prediction and assessing its reliability.

As has been well documented in the finance and data science literature, RBP extracts as much information from complex datasets as machine learning models but more efficiently and with full transparency.^22,24^ Our paper highlights RBP’s extraordinary transparency based on a dataset of self-reported impact scores for patients who have been treated with opioids.

RBP represents a novel contribution to the medical literature, grounded in rigorous concepts long used in finance, and offers a transparent alternative to conventional model-based prediction. Since it is model-free, it can be deployed without large training volumes and, importantly, can accommodate incomplete datasets by explicitly accounting for missing information rather than excluding affected cases. RBP also preserves informative extremes by avoiding routine outlier removal, thereby maximizing the utility of real-world clinical data while keeping the logic of each prediction auditable. Together, these features may yield more accurate and equitable decision support—helping clinicians and health systems better target interventions that are most likely to succeed, reduce avoidable harm, and ultimately conserve scarce resources and cost while minimizing deleterious effects.

## Data Availability

The data are proprietary and not publicly available; requests for access may be directed to Celeri Health, Inc. and are subject to a separate agreement.

## Contributions

CLR, DT, MK, RJY devised, wrote, and edited the manuscript. DT and MK performed the analysis. LL provided expertise, edited, and revised the manuscript.

## Data statement

This study is a secondary analysis of existing data from the PainRWD™ Science Cloud, provided by Celéri Health, Inc. under a data use agreement. The data are proprietary and not publicly available; requests for access may be directed to Celéri Health, Inc. and are subject to a separate agreement. PainRWD™ is a trademark of Celéri Health, Inc. Celéri Health, Inc. reviewed the manuscript for accuracy of data description and attribution. No new human data were collected, and no identifiable private information was accessed or analyzed. In accordance with U.S. federal regulations (45 CFR 46) and the principles of the Declaration of Helsinki, this secondary analysis does not constitute human subjects research and did not require institutional review board (IRB) approval.

## Acknowledgement of Affiliation and Funding

Christopher L. Robinson, MD, PhD, David Turkington, CFA, Linda Lee, MD, Mark Kritzman, MBA, CFA, and R. Jason Yong, MD all have a commercial interest in Cambridge Prediction Analytics. No funding was provided for the project.

## Independent Validation

Although these results highlight RBP’s distinctive features, independent empirical validation is needed to establish whether these features translate into superior predictive performance relative to alternative methods.

## Appendix Proof that Weighted Average Fit Equals R-squared for Linear Regression

Recall the definition of fit from Equation 6. We may rewrite fit equivalently and with full generality as the squared correlation between weights and outcomes:

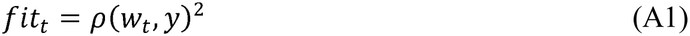

Let the prediction weights equal the linear regression equivalent weights from Equation 10, which we may write as a vector in terms of matrix notation with a vector of ones, 1_*N*_, and a matrix of predictive variable values, *X*.

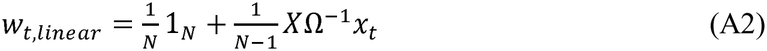

For notational simplicity, define the centered observation weights as 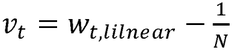 and the centered outcomes as 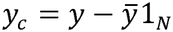. These definitions lead to the following two identities:

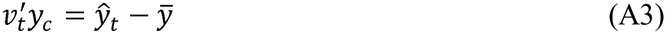

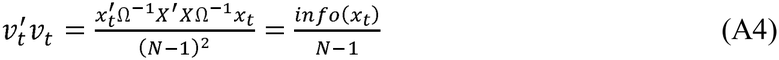

Equation A1 then becomes:

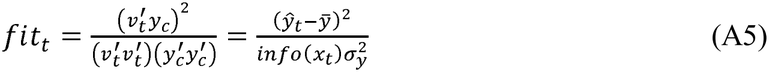

Define weights *a*_*t*_ for every prediction *t* in the observed sample based on the informativeness of each prediction task. These weights are nonnegative and sum to one owing to the properties of informativeness.

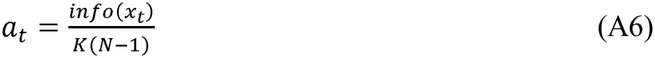

If we multiply fit by the number of predictive variables, *K*, and compute the weighted average across all prediction tasks, we find the result that it equals the classical R-squared statistic for linear regression:

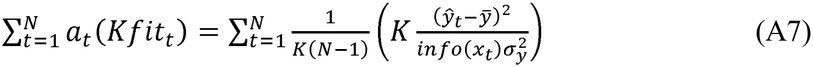

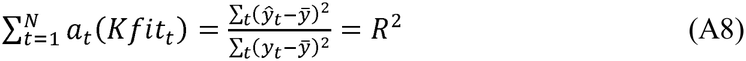

